# Emergency Medical Systemic frustration of Aggression and Violence in Conflict Encounters: An Evolved Grounded Theory

**DOI:** 10.1101/2023.07.24.23293095

**Authors:** Nigel Rees, Claire Hawkes, Lauren Williams, Julia Williams, Peter O’Meara

## Abstract

**Background:** Emergency Medical Services (EMS) staff worldwide have long been at risk of encountering violence and aggression (V&A) at work, including threats, verbal, physical, and sexual assault, and on rare occasions, fatalities occur. Exposure to V&A can result in stress, fear, and burnout. This is an international problem and EMS employers, trade unions and others are working towards tackling it. This paper reports the results of a qualitative study that aimed to explore protecting EMS Staff from aggression and violence in conflict encounters

**Methods:** This study took place in 2022 in one UK ambulance service covering a population of three million people. Individual, one-to-one semi-structured interviews were conducted with EMS staff via a Voice over Internet Protocol VoIP (VoIP). Data were analysed through Evolved Grounded theory methodology.

**Results:** Ten EMS staff were interviewed, and the following categories emerged: *Rusted, busted and inevitability of Violence & Aggression in EMS Environment, Tolerable or intolerable Violence & aggression in EMS, Gendered violence & aggression and Genderization in EMS, modifiable factors and harm reduction of Violence & aggression in EMS, Professional, ethical & clinical judgments of Violence & aggression in EMS, and Socio-cultural and system frustrations of Violence & aggression in EMS.* The Basic Socal Process (BSP) that emerged was one of *Emergency Medical Systemic frustration of Aggression and Violence in Conflict Encounters*.

**Conclusion:** V&A directed toward EMS staff is complex and our participants revealed how it has long been accepted and may be influenced by systemic frustration in EMS. Staff reported frustrations due to tiredness and in patients from delays in EMS response. Female EMS staff experienced sexual V&A by co-workers and patients, and these voices reverberate with female EMS staff internationally. Our study revealed how EMS has not effectively tackled V&A and many sociocultural constructs accepted within EMS. We call for the voices of our participants to be amplified internationally and for purposeful efforts to continue to be made in tackling this issue.

## Introduction

Emergency Medical Services (EMS) staff internationally experience violence and aggression (V&A) in their workplace (Yan et al 2013, Murray et al 2019, Rees & Whitfield 2005). This can involve verbal, physical and sexual V&A and fatal injury (Bigham et al 2014, Bernaldo-de-Quiros et al 2015, Maguire et al 2005, 2023). Experiencing such V&A can result in stress, fear, anxiety, emotional exhaustion, and burnout syndrome (Taylor et al 2016, Yoon et al 2016, Bernaldo-De-Quirós et al 2015).

Up to 90% of EMS personnel report V&A in their workplace (Corbett et al 1998, Suserud et al. 2002, Boyle et al 2007, Murray et al 2019) and is increasing (Maguire et al 2005). EMS systems internationally have attempted to tackle this problem through training, community education, policies and legislation, yet little is known of their effectiveness (Maguire 2018, Murray et al 2019, Rees et al 2021a, Spelten et al 2022). In Wales (UK) this has involved policy initiatives, such as the Obligatory responses to violence in healthcare (2018), changes in legislation including the Assaults on Emergency Workers (Offences) Act (2018) and media campaigns JESG #WithUsNotAgainst Us. Our team has recently described this multi-agency approach and the complex clinical, societal and legislative issues and we have developed a program of research and innovation which aims to explore potential strategies for protecting EMS staff from aggression and violence in conflict encounters (Rees et al 2021a). This paper reports the results of interviews with EMS staff and presents the results in a constructed evolved grounded theory.

## Methodology

We used Evolved Grounded Theory (GT) methodology (Strauss & Corbin 1998, Charmaz 2006) which is an inductive, theory-discovery methodology which sits within the constructivist paradigm of inquiry where our research team are passionate participants who facilitate a multi-voice construction of protecting EMS Staff from aggression and violence in conflict encounters (Lincoln & Guba 2005 p. 196). This included experienced academic research paramedics; their background and insider positions are important, as theoretical constructions are *interpretations made by the researchers and researched from respective given perspectives* (Strauss & Corbin, 1994, p. 279). We argue this perspective adds a richness of insight, awareness, and ability to bring meaning to the data (Strauss and Corbin, 1990). We do however recognise the potential for preconceived ideas and previous encounters to influence the construction of theory and have included techniques to assure trustworthiness.

## Methods

### Sample selection

Data collection was conducted between 2022-2023 in one UK ambulance service, covering a population of three million people. An electronic poster requesting volunteers was distributed throughout the whole ambulance service. Responses to were collated and participants selected using theoretical sampling based on characteristics identified by a review of the literature. Following the poster call, study participants were contacted by email, consented and invited for interview.

### Ethics

The study was approved by the Health Research Authority (HRA)/ Health Care Research Wales (HCRW) Integrated Research Assessment System (IRAS ID 313346). Participants were provided with a pack by email including and information sheet and consent form which they completed and returned and were also consented verbally during the interview. All data were collected and stored in accordance with GDPR (2018) regulations. Due to the sensitive subject matter, a range of support was available to participants.

## Data Collection

### Interviews

Individual, one-to-one semi-structured interviews were conducted by Microsoft TEAMS™ which may be considered a Voice over Internet Protocol VoIP (VoIP) and has been successfully used previously by our team (Rees et al 2021b). Interviews were one-hour duration using a semi-structured interview guide, video recorded with notes taken and transcribed verbatim.

### Data analysis

Analysis followed Strauss and Corbin’s (1998) three levels of open, axial and selective coding which involved “*explication of the story line*” (Strauss & Corbin 1998, p. 148) and identifying the Basic Social Process (BSP) around which all other categories revolve and development of an Evolved Grounded Theory. We have successfully used this methodology previously with paramedics (Rees et al 2018, 2021).

Transparency and trustworthiness was observed by a second researcher independently reviewing the coding framework and then discussing this framework with the primary researcher. Reflective notes were made, and member-checking’ techniques were used where the primary researcher returned to study participants to check their response and the construction the EGTM.

## Results

Ten EMS staff were interviewed. Figure 1 presents demographic data of EMS staff included in the study.

**Figure 1.**
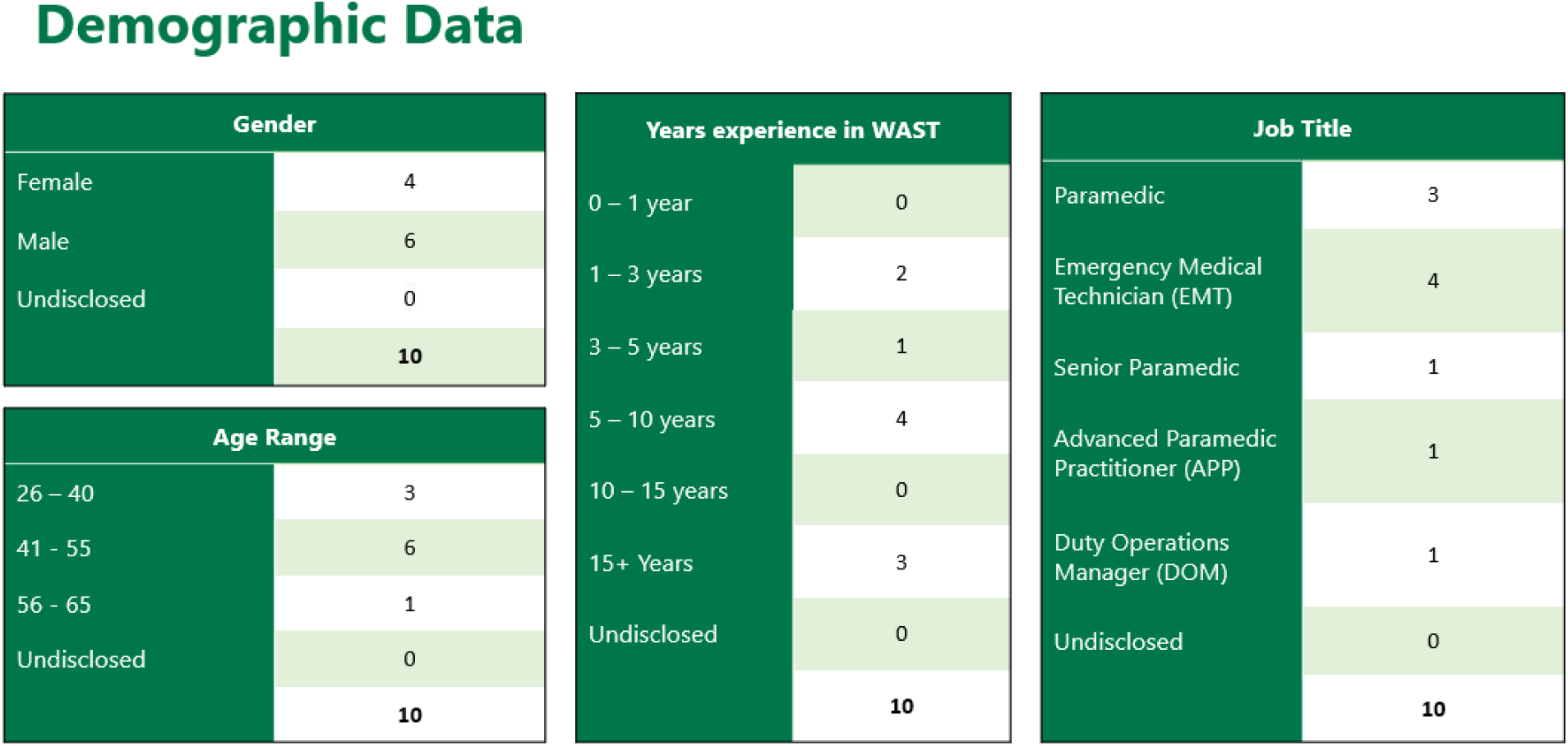
Demographic data or EMS staff included in the study.

**Figure 2.**
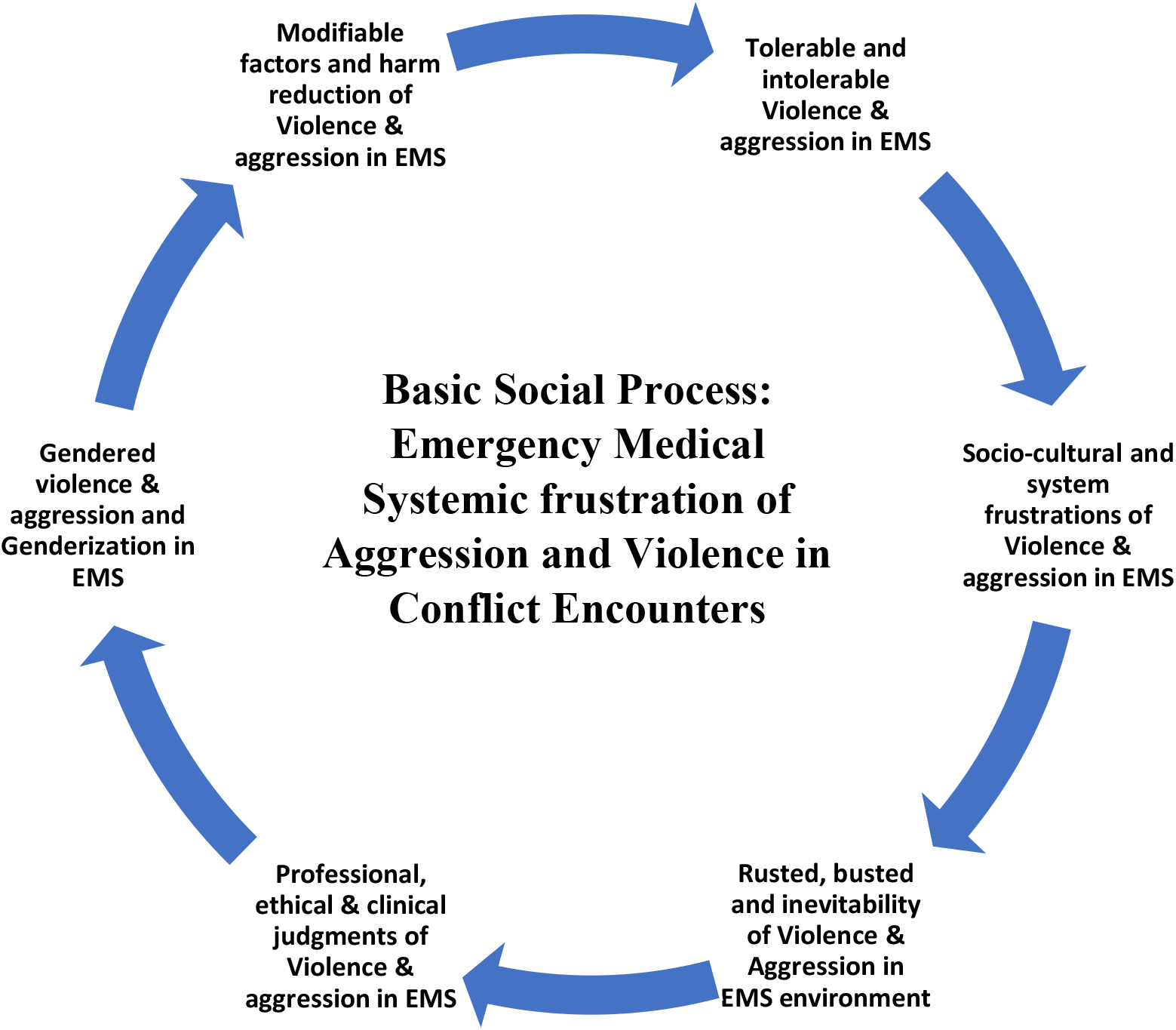
Evolved Grounded theory (EGT) of Emergency Medical Systemic frustration of Aggression and Violence in Conflict hEncounters

The following six categories emerged: *Rusted, busted and inevitability of Violence & Aggression in EMS Environment, Tolerable or intolerable Violence & aggression in EMS, Gendered violence & aggression and Genderization in EMS, modifiable factors and harm reduction of Violence & aggression in EMS, Professional, ethical & clinical judgments of Violence & aggression in EMS, and Socio-cultural and system frustrations of Violence & aggression in EMS*

The BSP which emerged was one of *Emergency Medical Systemic frustration of Aggression and Violence in Conflict Encounters* where patients EMS staff and others experience frustrations which can sometimes result in V&A.

### Rusted, busted and inevitability of Violence & Aggression in EMS Environment

Our participants reported how most people they encounter do not present a threat of V&A to them and talked about the trusted context of care they provide, involving intimate contact and invasive procedures. They told how this context inevitably made them vulnerable to V&A and its physical and emotional consequences and they all reported times of being scared and that V&A was increasing. Powerful experiences were shared of a wide range of V&A including verbal abuse, threats, sexual and physical assault with body fluids such as vomit and spitting, and weapons such as pens, screwdrivers, and knives. Participant 9 shared a terrifying assault on him by a housebound elderly man:

> *“his right arm came around, grabbed the carving knife, he brought it right around, just underneath my chin there, and his words to me was listen don’t fucking start” (P9)*

V&A was seen as just part of their job, but that it would not be accepted in other workplaces:

> *“Although we are frontline workers, you wouldn’t expect it in an office job, you wouldn’t expect it in a college, you wouldn’t expect it as a teacher, so it’s unacceptable. Is it understandable? Very much so, and you can understand the processes”* (P2.)

Our participants talked about the inevitability of V&A within the EMS environment due to the context of dealing with people who may be intoxicated with drugs and alcohol, those with impaired mental capacity such as the elderly and medically ill, or responding to people who are frightened, in pain and frustrated. Participant 2 said:

> *“I think there is an acceptance at some point you will be assaulted”* (P2.)

While participant 3 said:

> *“sometimes it comes with the job that we’re doing. If you look at that drunk, we’re picking up, if you look at that family, that sort of whatever the road accident of nobody intoxicated but the adrenaline is so high that they’re abusive and verbally abusive, we have to de-escalate them scenarios”* (P3)

They also reported being tired and stressed and talked about their exasperation with hospital hand-over delays.

> *“with waiting times, and hospitals, you know the way things are. That is stressful. You know the common thing you hear is we didn’t train for this, we didn’t train for waiting for hours on end but it’s become part of the job that we do…. it is frustrating when you’re sitting there but you cant allow that frustration to overspill into the way that you speak to and treat patients, you’ve got to be able to sort of deal with that frustration and some people just don’t seem to be able to” (P8)*

And

> “*The crews that we’re sending out potentially are more burnt out, stressed? Fed up, or what’s the other one I’ve learned recently? rusted out…. So instead of doing the job you’re sat outside hospital for 12 hours babysitting a patient, so you’re sort of rusting like*” (P2.)

All our participants recognised the potential for EMS staff to contribute to the V&A and how a minority of staff seemed to receive more V&A than other colleagues:

> *“I think sometimes, some crews through their own frustration sometimes don’t help themselves. I’m not saying everyone deserves what happens, not at all. However, I have been with people and you’re thinking, if you speak to them like that, you’re just going to wind them up”* (P7)

### Tolerable or intolerable Violence & aggression in EMS

Participants talked about V&A directed towards them by patients, bystanders, relatives, friends, and co-workers. Whilst they felt it was never acceptable, they acknowledged it could be understandable in some situations. They reported how a wide range of medical conditions could present with V&A, such as dementia, mental problems, head injury, or diabetes. They were understanding and tolerant towards such patients and talked of the importance of not missing such conditions requiring prompt treatment. They did however acknowledge that it should never be acceptable to face such V&A within the workplace, as articulated by participant 9:

> *“Understandable, to a certain extent, so like you know, we have staff you know, getting assaulted by patients with dementia or you know, the elderly patient has got various infections which affect them sort of mentally, I understand it a little bit more, is it acceptable? No not to me, it isn’t acceptable because it shouldn’t be acceptable for anybody to go out, do their job, and get abused and assaulted in the process of doing your work. So, I can understand it, but as far as being accepted?”* (P9)

Participants were less accepting of V&A from those intoxicated with drugs and alcohol which they appeared to find intolerable. This intolerance seemed to emanate from the deliberate nature of V&A by those who choose to become intoxicated from alcohol or drug taking as opposed to medical conditions deemed to be less within their control. Participant 2 said:

> *“it’s not the patient wanting to hurt me, it’s the patient or the illness. Whereas…the sort of under the influence Friday night drunk – It could be the alcohol talking, not him or her, but they’ve chosen to potentially get into that state… there’s a choice that there’s a series of choices that have led them to their presentation, whereas things like dementia, delirium, UTIs, that kind of thing. I wouldn’t say there’s a conscious “I’m going to get a UTI so I can scrap”.* (P2)

### Gendered violence & aggression and Genderization in EMS

All participants reported awareness of sexual and gender-related V&A in EMS. This ranged from sexism and inappropriate language to sexual assaults by patients and co-workers resulting in dismissal. Four of the participants had experienced of V&A and sexual assault by co-workers; all victims were women, and all perpetrators men. They reported experiences including inappropriate and sexual comments, groping of breasts and buttocks, and men revealing themselves. Participant 6 talked about her experiences with a patient:

> *“we were, trying to turn him onto the bed umm, and he groped me and then again, we were lifting that patient from a chair and he was in a position to grope me again”* (P6)

And

> *“one wasn’t a patient, one was a crew mate who sexually assaulted me and then consequently in the two months after, was two elderly patients who didn’t have any neurodegenerative diseases, so yeah that was fun, they were both sexual assaults as well”* (P6)

Participants talked about a perceived acceptance of this behaviour within EMS. Participant 9 shared her experiences of how this behaviour in patients had long been tolerated, telling how:

> “*it just used to be the oh they’re drunk or things like that, or you know, they don’t really know what they’re doing, which is questionable*” (P9)

While participant 8 shared her experiences of tackling such behaviour in patients:

> *“I’ve had to talk to the patient and say, that’s completely unacceptable and its not appropriate to be talking to my colleague in that kind of way or to be making that kind of suggestion*” (P8)

Such acceptance of sexual and gender-related V&A was also shared by participants in relation to EMS colleagues. Those who had been sexually assaulted by co workers told of their discomfort in working with their assailants prior to the assault and how their behaviour was known to other colleagues and tolerated. Participant 7 powerfully shared her experiences and escalation of this behaviour:

> *“I never used to like working with him it’s just my little spider senses, I wasn’t comfortable, and he used to say things and people just took it, that, that was just him. That was just him being perverted and – so therefore I didn’t like to speak up”….“he probably felt he got away with it until this one night, thought he could, reveal himself, umm, on a night shift, which totally threw me. Umm, and was totally unwarranted, totally unacceptable, umm, but put me in a – I didn’t know what to do”* (P7).

And

> *“So yeah, but I think if that wasn’t accepted, I could have spoken up earlier and said I don’t like this and unfortunately because of my background I didn’t have the confidence to say you know, what do you think you’re doing? I was like oh go away, go away and inside panicking thinking, this is not right, I don’t feel comfortable, but because that’s the way of the ambulance service that those things were oh it’s fine, it’s banter, it’s not quite clear as what’s banter, and I wasn’t comfortable but”* (P7)

And

> *“I think it’s just like the line isn’t it? Between banter and being sexually abusive, sort of unacceptable. It’s just that line that we seem to, as I said for me, it was – everybody saw him as, oh he’s a pervert”* (P7)

Participants exhibited an element of genderisation; for instance when referring to older people presenting with illness such as dementia the perpetrator was referred to in the feminine form, whereas younger assailants intoxicated with drugs and alcohol were referred to in a male form as evident in comments by participant 2:

> *“I think there is an acceptance at some point you will be assaulted, and I’d hope from my point of view, if I am assaulted again, I hope it’s by little 86 year old Doris in a nursing home as opposed to coked up Dave in the street. Because, although they are the same one paper, they’re really not.”* (P2). [NB names and ages are not those of real people]

### Modifiable factors and harm reduction of Violence & aggression in EMS

Many modifiable factors and opportunities for harm reduction were presented by our participants. They talked about the need for training, opportunities to intervene early in the call cycle, communication, media, meal breaks, and efforts to address tiredness and reduce hospital handover delays. They gave examples of technologies being used to protect staff and address V&A, including flagging addresses of perpetrators, panic buttons on radios, stab vests, spit masks and spit tests to protect and gather evidence for prosecutions, body-worn cameras (BWC’s) and vehicle cameras to prevent V&A and record encounters for evidence and training.

Participant 4 said:

> *“ [control] put a red flag on the address so that any crews that were called there, would know that a patient has branded a weapon at us, umm, and that they wouldn’t send solo’s as well”* (P4)

They did however report disappointment in having to introduce such things into EMS, and the potential consequences. BWC’s for instance were seen as an invasion of privacy which may escalate V&A. Participants also talked of their experiences with simple improvements such as their frustrations at reporting incidents of V&A, how it could be a waste of time, and that they rarely had feedback. Participant 3 said:

> *“for everyone that’s reported there’s probably 10 that haven’t been reported”* (P3)

And

> “*the amount of statements I’ve done with police is ridiculous and nothing has happened*” (P6)

Participants provided sensitive accounts of the emotional burden of reporting, and their attachment to this process which extended into their personal lives and influenced their recovery. Participant 6 told of her ongoing distress and unresolved issues following an attack and how she received a call whilst shopping with her infant child:

> *“I was walking around a supermarket with him [infant child], and I had a call saying “oh the person you reported has died, so we’re not going to take it any further” so I was like ok fair enough”* (P6)

For some there was a wish to see justice being served, to punish perpetrators and prevent future encounters, but many were doubtful this was happening, as told by participant 2:

> *“the legal system itself is struggling with these cases because you know what’s the deterrent? Probably – I probably get paid £50 in 10 pence instalments, as a victim surcharge, they do some community service. Where you know, where’s the deterrent in that? There isn’t any”* (P2)

Others reported the loss of control by their perpetrators at the time of the incidents and the remorse they would feel when not in that state. This presented a dilemma for some staff where perpetrators of V&A were also viewed as patients with complex needs and vulnerabilities. Participant 7 said:

> *“I had the choice of taking it further to possibly prosecute or mediate with the patient, and I chose mediation, because she had a child who was sort of, palliative care, and I felt it wouldn’t have been in her benefit or mine really”…. “she took that substance, rightly or wrongly just to cope and just to have that moment of escapism and I felt that if I could chat to her and just let her know the consequences of her actions,* (P7)

Participant 7 also reported the following powerful and empathetic exchange between her and her perpetrator:

> *“we had a chat because we couldn’t meet up due to COVID and she was just crying and crying and so apologetic and to me, I felt that, I could sort of put myself in her position and not sympathise but just sort of maybe understand a bit”* (P7)

EMS staff in our study told of the importance of being listened to and the role of policies and groups such as the National Health and Safety Group, colleagues, managers, and their trade unions. Participant 7 shared how:

> *“its just frustrating to me to see that they, that management could have such a better, more supportive workforce if only they would just sort of – just try, looking into how crews are supported. And this is it, they keep saying why are so many crews off, I could tell them why lots of crews are off, they’ve just had enough”* (P7)

Whilst it was recognized that there was still a lot of work to do, many staff reported the importance of supporting them through this process and its role in their recovery. Participant 6 told of the support he had received:

> *“For PTSD, which I still do struggle with now, but umm, it was brilliant, really good and it really helped, hence why I can talk about it, because I wouldn’t have been able to previously, so really good stuff”* (P6)

Many spoke of the lasting emotional and psychological effect the V&A encounter had on them along with the subsequent investigations. Participant 7 told of the lengthy processes she endured following sexual assault by a co-worker and how:

> *“the [The Trust] has been amazing. [NAME], do you know [NAME]? She’s in [DEPARTMENT] oh my god, she saved my life. She was amazing, amazing. Umm, and they sent me for EMDR counselling. Because obviously what he did then opened a can of worms, so, you know, I had my days but, on the whole, I was totally supported by work. Work were absolutely amazing, amazing, so I was lucky” (P7)*

### Professional, ethical & clinical judgments of Violence & aggression in EMS

Participants talked about the professional, ethical & clinical judgments of V&A in EMS. They gave examples of the challenges faced and judgments they had to make around the assessment and management of patients presenting with conditions such as head injuries and diabetes where V&A could occur.

> *“there’s so many different conditions out there now where violence can be a symptom…. hypos, head injuries, or any other metabolic disorder, so it’s something that we’ve always got to be very mindful of”* (P4)

They presented many difficult scenarios, navigating such risks and duty to care. Paramedic 8 said:

> “*if they said anything like, well you know, I’m going to kill myself if you kick me off the ambulance or whatever, well, are they? Don’t know. Can you take that risk? Not really”.* (P8)

They shared their vulnerability to V&A when weighing up risks to patients of not receiving their care as discussed by paramedic 9:

> *“we could request that we stand off until somebody is there to help us, but you know, at the end of the day you’re being told somebody is dead and you know that’s what we’re all trained to do is for those situations so I suppose, you know, we do sort of put ourselves in vulnerable positions at certain times”* (P9)

### Socio-cultural and system frustrations of Violence & aggression in EMS

Our participants gave examples of socio-cultural and system related sources of frustration, influencing V&A, including the role of alcohol and drugs, responding to disturbances, patients and their relatives being scared and in pain. They also talked about communication challenges, all told how the attitudes of EMS colleagues could incite V&A where they felt the same EMS staff got assaulted due to their own poor attitudes and communication.

> *“The public are not always at fault… staff are not wholly innocent”* (P1.)

And

> “*I’m not innocent, I’m not, but if we go in there a bit bolshy, you know, when they’re fragile anyway they, you know… but people are – staff are overtired. They haven’t been fed, they haven’t been watered, whatever is going on or they’re burnt out, or they’re stressed out, if they’re going to go to somebody’s house who’s been waiting over the – over the time and the relatives are a little bit narky only a tiny bit that could be enough to – to give them attitude, you know?*” (P1.)

And

> *“because they’ve got their own frustrations, their own, they’re upset with work, they don’t feel supported, they don’t feel valued and all this, they then hold onto that frustration and it can, not always, but it can come across with their patients then”* (P7)

All our participants cited delays in ambulance responses as a source of frustration to patients and their family members which could result in V&A. They described how they were often the first person a caller encounters and their efforts to de-escalate the situation:

> “*something that we’re seeing more and more of is patients saying we’ve waited so long for you, I can understand why the public would be upset if they’ve had to wait 6,8 maybe even longer hours, for what they see as their emergency, when we walk through the door, I can see why people would be potentially upset at that, that it has taken so long umm*” (P.4).

And

> *“when they sat waiting and waiting [for an ambulance] and they’re getting more and more worked up, for whatever reason, they might have taken drugs, they might, you know, they’re desperate aren’t they? So, when we knock the door then, they’re like – they just erupt and we’re gonna get it both barrels”* (P1)

They described efforts of tackling V&A such as calling for help from colleagues and the police to detain and prosecute perpetrators. Participants were mostly aware of recent legislative changes, and whilst many could not name the Assaults on Emergency Workers Act (2018), they supported raising awareness of this issue and this legislation with the public. They shared personal experiences which resulted in prosecutions,

> “*that is what that lady was prosecuted under when she spat at me and the police officer. I understand that it allows for harsher sentencing than potentially if it had been a member of the public towards another member of the public so I really, think its an excellent element of law”* (P4)

Some participants did however report doubts over the effectiveness of the legislation within the context and that it may not be effective in deterring or preventing V&A as:

> *“No amount of Acts is going to change peoples attitude, if they have that inclination to act violently and aggressively towards somebody, either because they feel threatened or they’re being asked to do something they don’t want to do, they are just going to act like that”* (P8)

These doubts were also accompanied by a degree of cynicism, that such legislation and efforts would be effective, and were politically motivated to appear to be doing something about this issue as shared by participant 2:

> “*I fail to see how, that – that might be my warped opinion because I have seen some political commentators and legal commentators talking about it… then their core message is that it’s a political statement as opposed to a real one… It’s like trying to stick a plaster over the whole of the Titanic, it’s a bit of a holding point as opposed to a meaningful change”* (P2)

The role of such legislation and communication in preventing V&A was questioned by participants:

> “*if you’re in a state of mind where you can remember the law, you’re probably not going to assault somebody in the first place or act in a manner that is violent or aggressive, so I suppose, it’s probably not going to act as a deterrent, however what it will do is get those potentially dangerous people off the streets, isn’t it”* (P4)

### Evolved Grounded Theory: Emergency Medical Systemic frustration of Aggression and Violence in Conflict Encounters

*Emergency Medical Systemic frustration* emerged as the BSP of Aggression and Violence in Conflict Encounters within EMS. We draw on the frustration–aggression hypothesis first proposed by Dollard et al (1939) who states: “*the occurrence of aggressive behaviour always presupposes the existence of frustration*” (p. 1). Many actual experiences of frustration were reported by participants in our study, including EMS staff, patients, bystanders, and others which spanned all the categories. Dollard et al. (1939) define frustration not as an emotional experience, as in these cases, but as “*an interference with the occurrence of an instigated goal-response*” (Dollard et al., 1939, p. 7). Frustration is therefore viewed as an event or action that complicates the accomplishment of a task, or as Berkowitz (1989) proposed in his reformulation of this hypothesis, that: “*frustrations are aversive events and generate aggressive inclinations only to the extent that they produce negative affect*” (p. 71). Whilst there may be other sources of aggression, it is widely acknowledged that frustrations generate negative affect, which, in turn, can lead to or increase aggression (Berkowitz, 1989, Marcus-Newhall, Pedersen, Carlson, & Miller, 2000).

## Discussion

The range of V&A reported in our study is also commonly encountered by other EMS staff internationally, who experience verbal abuse (67%-99%), intimidation (41%-80%), physical assault (26%-69%), sexual harassment (14%-61%), and sexual assault (3%-4%) (Bigham et al 2014, Boyle et al 2007, Savoy et al Mausz et al 2021, Newbury-Birch et al 2017, Mausz & Johnston, 2019). Our study also reflects previous research where EMS staff appear to accept V&A is just part of the job involving a complex interplay of contextual and systemic issues within their working culture and environment (Mausz & Johnston 2019, Boyle 2007).

We found some forms of V&A were tolerable, such as in older patients presenting with disease such as dementia which has also been reported by others (Byon, et al 2020). Conversely, our participants conveyed intolerance to V&A involving drugs and alcohol due to its deliberate nature, which may be subject to choice, rather than in injury or illnesses which may be less under less control. Despite this, drugs and alcohol presentations have high mortality and EMS staff have long played a crucial role in their care, often being the first and only point of medical contact (McCann et al 2018, McDermott & Collins 2012, Bolster et al 2023).

All participants highlighted how behaviour of EMS staff may contribute to V&A which may have many influences, such as individual personality, presupposition to V&A, limited communications skills, and training. Systemic factors and frustrations in EMS were also forwarded, such as long hospital delays, busyness, tiredness and stress in themselves and colleagues. The *Rusted and Busted* metaphor presents these issues and reflects assertions of Bolster et al (2023) that the role of EMS staff in drugs and alcohol crisis may be complicated by increasing burnout, helplessness, and moral distress, and this once rewarding patient encounter has evolved into one which may leave both parties (the patient and the paramedic) feeling dissatisfied. Bolster et al (2023) forwards the notion of such cumulative stress as potential contributory factors through the multiple studies identified in their review, revealing that paramedic students have the lowest levels of empathy for people who use drugs than any other patient population, and some view them as unworthy of medical treatment, and a burden on the medical system (Williams et al 2012, Williams et al 2015a, Williams et al 2015b, Kruis et al 2021, Pagano et al 2019, Kus et al 2018, Williams et al 2015 c). We have also previously reported how V&A may be influenced by compassion fatigue, which can occur when practitioners become emotionally exhausted and lose the ability to respond empathically to their patients (Rees et al 2018, Gillespie et al 2003).

Many of the frustrations raised by EMS staff relate to wider system issues which may indeed be influencing such V&A. We have previously reported similar systemic issues in EMS, including training, tiredness, appropriate care options and safe alternatives to ED for people with mental health problems and alcohol intoxication (Rees et al 2018). Decision-making of EMS staff in a context of risk has also been reported to be usual in this context; especially when faced with patients who present with V&A, lack mental capacity, or appear intoxicated with underlying medical problems or injuries (Rees et al 2018).

This frustrated environment where V&A is so prevalent may be creating conditions for, and reinforcing, the potential of further V&A. Work frustration is the negative emotion resulting from unsatisfied needs and motivation due to the obstacles and interference experienced by an individual in a workplace (Fox & Spector, 1999). The psychological explanation of frustration aggression is not without criticism as frustration does not always lead to aggression. People can indeed decide to be aggressive without being frustrated. The theory excessively focuses on the individual’s internal mechanism, but such individual action occurs within a wider social context, and hence we argue this frustration is not solely observed in the individual, but rather, the frustrated wider EMS system. Such system frustrations and reports by EMS staff reflect the challenging factors related to this issue, and previous research reports between 52 - 81.6% of cases of V&A on EMS staff is associated with mental behavioural disorders (MBD) or significant mental illness, 47% of which requiring psychiatric admission (79%, involuntarily), 11% have a history of violence, 24% are repeat perpetrator (93.5% associated with a MBD), 17% - 42.4% involve alcohol, psychoactive substance or illicit drugs (Knott et all 2005, Thomas et al 2022).

The frustration-aggression theory, suggests that V&A can occur as a result of the disparity between what is sought or desired and what is obtainable in reality (Dollard et al. 1939;; Amenta 2005). Our participants all cited delays in ambulance response as a major factor in V&A, and such delay has previously been reported as a predisposing factor for V&A in other emergency settings (Corbett et al 1998, Bernaldo-De-Quiros et al 2015, Rahmani et al 2012, Koohestani et al 2012, Sheikh-Bardsiri et al 2013). Indeed, delays and other complicating contributors, negatively influence a strayed relationship between people who use drugs, EMS systems and its staff (Mamdani et al 2022, Bolster 2023). Patients may be in pain or injured, where themselves, relatives or bystanders may become frustrated when the desired goal of receiving care is not attainable, provoking an emotional response towards perceived authorities standing in the way of his achieving or attaining his desire. Waiting times may be seen as a source of frustration and irritation (Spelten et al 2020, pich et al 2010, Kennedy et al 2005), indeed, Morken et al (2015) reported how ED Nurse participants perceived episodes of V&A to be the result of a mismatch between patient expectations and the actual service.

All participants in our study recognised the significant volume of sexual related V&A, which is also reflected in the literature with14%-61% of staff reporting assault and harassment and 3%-4% sexual assault (Bigham et al 2014, Boyle et al 2007, Savoy et al Mausz et al 2021, Newbury-Birch et al 2017, Mausz & Johnston, 2019). None of our male participants reported personal experiences of this but recognised its presence. All female participants however, except one, revealed powerful and disturbing experiences of sexual assault by patients and co-workers; they wanted to speak up and have their stories heard. Whilst both males and females are victims of sexual harassment and assault in EMS, it is experienced more in females than males (34.2% against 9.2%) (Boyle et al 2007, Savoy et al 2021, Shabanikiya, et al 2021), this picture is reported in other professions (Akhter et al 2019, Ansoleaga et al 2019, Chappell et al 200, 2006, Rotundo et al 2001). Within our study, however, many of these were perpetrated by EMS co-workers, which has also been reported elsewhere (Boyle et al 2007).

McFarlane et al (2022, 2021, 2020) reported how female paramedics experience everyday sexism, gender discrimination and sexual harassment in the workplace which influenced how they practiced, and negatively impacted on career progression and well-being. The Victorian Equal Opportunity and Human Rights Commission (2022) recently conducted an inquiry into the culture at Ambulance Victoria (AV) where 83% of staff reported sexually suggestive comments or jokes and 10% indicated they had experienced requests or pressure for sex or other sexual acts, 3.6% had been subjected to actual or attempted rape or sexual assault. AV is a jurisdictional ambulance service in Australia servicing a population of almost 7 million people across urban and rural settings that is, comparable to Wales. AV was recognised in this report to be an increasingly tired and frustrated community, but this may reflect EMS staff experiences internationally as Mausz & Johnston, (2019) also found 61.5% of EMS staff report sexual harassment, and 13.8% sexual assault.

The Victorian Equal Opportunity and Human Rights Commission (2022) reflected an unawareness in senior leaders of this culture of sexism in EMS. The National Guardian’s Office (UK) (2023) also recently conducted a ‘*Speak Up’* review of NHS ambulance trusts in England and found reports of sexism and a culture that did not support workers to speak up. A ‘macho’ culture has also been suggested to exist within UK ambulance trusts is leading to widespread abuse of female staff (HSJ 2021). Whist such sexism and sexual abuse is reflected in the research literature (Hanna-Osborne 2022, Baranowski, L. and Armour, R., 2020 McFarlane et al 2022, 2021, 2020), few studies have explored co-worker sexual violence. Hanna-Osborne (2022) found that out of the 30 participants, 25 had experienced sex-based harassment from male colleagues. Most commonly this took the form of gender harassment such as comments and jokes designed to belittle and demean women based on their gender. Boyle et al (2007) however reported that a significant number of paramedics experience sexual harassment/assault in the workplace by work colleagues, and participants in Hanna-Osborne (2022) experienced sexualised forms of harassment, including unwelcome sexual attention and propositions. They expressed reluctance to report the behaviour through organisational channels because of the perceived futility of doing so and the potential for reprisals and career repercussions. The preferred responses to harassment were informal, and included avoidance, humour, direct appeals, and work withdrawal.

The sexualised and sexist culture within UK Ambulance services is not new, and has been reported over many decades, including accounts of serious incidents, exposure to pornographic material, physical propositioning, and sexual assault (Devon Live, Norther echo 2015 Loweth 2013, HSJ 2021, Mausz & Johnston, 2019). Female EMS staff have talked about ‘sexual predators’ among male colleagues who ‘groomed students’ for sexualised ends (Guardian 2017) which also reflects Sheen et al. (2012) who found that 16% of paramedic students were exposed to unwanted sexual behaviour.

We also noted genderisation when referring to older people presenting with illness such as dementia were the perpetrator was referred to in the feminine form, whereas younger assailants intoxicated with drugs and alcohol presented as males. Whilst further work is needed to establish the accuracy of such perceptions, this may relate to wider societal gender norms and stereotypes which restrict individuals to gender identities and expressions that conform to the male-female binary. Such genderization is evident across wider society (Karniol, 2011, McDowell, 2021, Shaw-Garlock, 2014, Van Driel et al., 2018). Early work of Broverman et al (1970) suggests agency and communion as core dimensions of gender stereotypes gender. Agency incorporates traits such as competence, instrumentality, and independence; communion encompasses expressivity, warmth, and concern with the welfare of others. Such attitudes may however reflect the cultural problems above and machismo, which is a sociocultural construction where men are characterized as aggressive, independent, and dominant, and women are characterized as weak, dependent, and submissive (Pérez-Martínez et al 2021).

Mixed opinions were shared of the potential impact of legislation, ranging from support to doubts of its effectiveness within the context, and that it may not prevent V&A as perpetrators are often incapacitated or intoxicated during the V&A. This notion is also supported by Eburn, and Townsend (2018) who propose that in most people who commit V&A on EMS staff, such legislation will unlikely be at the forefront of their minds, and punishment after the event shows that the desire for protection hasn’t worked. As in our study, Eburn, and Townsend (2018) highlight how the very people who need EMS assistance are those suffering mental health crises, intoxication, and injury, and such people are unlikely to make rational decisions at that time. Eburn, and Townsend (2018) argue that punitive sentences such as jail will not decrease the risk of, or actual events of V&A, nor will it address the causes or reduce the risk of future offending.

Our participants reflected on the trusted context of their care, involving intimate contact and invasive procedures also highlighted by Eburn, and Townsend (2018) who argue this trust may be eroded if people contact EMS for unconscious friends or family members to be prosecuted or jailed if they become agitated or violent. Communications and media reports in Wales highlight doubling of maximum prison sentences under the Assault on Emergency Workers (Offences) Act (WAST 2022) which may be seen as a deterrent towards such V&A, but as was suggested in our study, Eburn, and Townsend (2018) argues that such legislation may be more politically motivated, for Governments to appear to be doing something about this issue. Some of our participants reported disappointment at low sentences, which again may reflect Eburn, and Townsend (2018) assertions (with examples) that despite Government rhetoric around mandatory sentences, all cases are different, judged on their own facts, by magistrates and judges not governments or legislators. Eburn, and Townsend (2018) highlights how reports focus on maximum sentences, which allow for lesser sentence in special circumstances which often includes offenders affected by drugs or alcohol at the time.

People who use drugs or alcohol often feature in media reports, but not as special circumstances, rather, in pejorative and demeaning language such as “drug addict,” “drug-addled stupor” and aligned with “thugs, perverts, drug dealers and other criminals” (Rees et al 2021a). This is not consistent with the professional and considered manner of our respondents when talking about these patients groups. Such pluralistic media rhetoric of villains being locked up for assaulting heroic EMS staff belies the complexity of this issue, may ultimately be unhelpful in tackling it, and rather, have the opposite effect of marginalising these patient groups further, eroding their trust in EMS staff and increasing the vulnerability of them and EMS staff rather than making things safer (Eburn, and Townsend 2018, Bolster et al 2023, Rees et al 2021a, NIDA 2021 Volkow et al 2021, Wagner et al 2019). The language used to describe the drugs and alcohol taking population therefore matters, and EMS staff and their leaders should therefore receive education in this area which has been shown to result in significantly less stigmatization of this population (Kruis et al 2021). Such education should start at the point of entry such as University and induction, then continue to be reinforced throughout their career through case based discussion, reflection and clinical supervision.

EMS leaders and staff therefore have a difficult balance to strike in protecting EMS staff when considering barriers of harm reduction, such as the willingness to activate EMS drugs and alcohol and decreasing police presence to reduce on-scene tension bred from longstanding negative from intergenerational trauma, criminalisation and stigma (Bolster et al 2023) as well as the opportunities, for post-drug poisoning therapeutics can be viewed as life-saving (Wagner et al 2019

Many modifiable factors where suggested, some of which involved technological innovations such as stab vests, spit tests and flagging addresses. Many have been introduced despite sparce evidence for their effectiveness. Participants for instance spoke of BWC technology which EMS systems are increasingly adopting at considerable financial cost, yet Bruton et al (2022) found no evidence that implementation of BWCs, alone or as part of a broader suite of initiatives, reduces the incidence of V&A towards paramedics and calls for further research.

Some modifiable factors however were cultural and systemic; for instance, some reported negative experiences and futility in reporting V&A, which reflects research which found only 40% of EMS staff report the incidents to service management and 21.3% to Police (Mausz et al 2021). Support from colleagues, unions managers and opportunities for to face meetings with the perpetrator seemed to be valued. One of our participants told of a formal meeting with her attacker as a form of restorative justice. Non-punitive intervention such as this aim to increase accountability for perpetrators at a societal level and have been trailed elsewhere which presents an opportunity to tackle V&A, but further prospective trials are needed in EMS (Thomas et al 2020).

In delayed EMS responses participants highlighted the role of the dispatchers keeping callers informed which may help resolving a mismatch between patient expectations and the service offered. Alerting EMS staff and involving the police where there is risk of V&A such as in cases involving severe mental health problems or intoxication with drugs flagging addresses validated risk.

Our participants suggested that efforts to tackle V&A such as legislation is like trying to stick a plaster over the whole of the Titanic… a bit of a holding point as opposed to a meaningful change. This reflects Eburn and Townsend’s (2018) assessment of the problem, that this sort of work is expensive, slow and unpopular in today’s rhetoric and being ‘tough on crime’. To protect paramedics the community needs to address offending before it happens fund education, mental health services, drug rehabilitation Eburn, and Townsend (2018)

## Strengths and limitations

Our study is limited in being conducted in one UK country and single EMS. Our sample also may reflect non-response bias where those interested in telling their stories may have been more likely to engage in the research. The research team included paramedics which again may be considered a potential bias, but we believe this adds added to the richness and ability to bring meaning to the constructed GT. We found consistency and agreement throughout analysis, member checking and previous studies which adds to the trustworthiness and transferability of our findings. We also recognise the significant limitation of this study and other V&A research in EMS, which does not include the voice of patients and perpetrators of V&A.

## Conclusion

Protecting EMS Staff from aggression and violence in conflict encounters is complex and multifaceted. Through interviews with EMS staff we have constructed an evolved Grounded Theory which proposes systemic frustration as a basic social process of Aggression and Violence in Conflict Encounters in EMS. We draw on the frustration– aggression hypothesis (Dollard 1939, Berkowitz 1989) which views frustration as an event or action that complicates the accomplishment of a task. EMS staff reported that V&A is inevitable within their working context and many issues emerged that complicate the accomplishment of the task of EMS staff providing care.

EMS staff provide care for people with medical problems which impair their mental capacity and may involve V&A yet is tolerated. This contrasts with V&A in those intoxicated with alcohol or drugs which they find intolerable, which can present significant professional, ethical, and clinical judgments and challenges including balancing risk to EMS staff against potential stigmatization barriers to care for these high risk groups. Views were mixed on media campaigns and legislation, and whilst most welcomed these initiatives, some saw them as a sticking plaster over more complex problems.

Systemic frustrations within EMS were revealed which may be modifiable in tackling V&A and included delays in ambulance response, tiredness, stress, lengthy hospital handover times, communication skills, and training shortfalls. Technological solutions were also forwarded such as BWV and other opportunities, such as validated risk decision tools for minimising frustration in delayed EMS response and dispatchers keeping callers informed. These opportunities are costly and require research to evaluate their effectiveness. The impact of V&A may also be minimised through policies that support the victim, and positive examples were provided of support from colleagues, managers and unions, along with restorative initiatives all of which require more research.

Gender featured prominently in interviews and may reflect outdated systemic gender norms and societal constructs of men characterised as aggressive and women submissive. Female EMS staff told disturbing stories of sexual V&A by co-workers and patients, and whilst some involved predatory criminals within EMS as in wider society, the voices of our participants reverberate with those of other female EMS staff internationally who face sexual V&A systemic within EMS culture. The frustrated EMS metaphor reflects a system that has not tackled these sociocultural constructs accepted within EMS and we call for these voices to be amplified internationally and purposeful effort made to tackle this issue.

## Data Availability

All relevant data are within the paper and its Supporting Information files.

